# ADVANCING PARKINSON’S DISEASE RESEARCH IN CANADA: THE CANADIAN OPEN PARKINSON NETWORK (C-OPN) COHORT

**DOI:** 10.1101/2024.05.10.24307196

**Authors:** Marisa Cressatti, Gabriel D. Pinilla-Monsalve, Mathieu Blais, Catherine P. Normandeau, Clotilde Degroot, Iris Kathol, Sarah Bogard, Anna Bendas, Richard Camicioli, Nicolas Dupré, Ziv Gan-Or, David A. Grimes, Lorraine V. Kalia, Penny A. MacDonald, Martin J. McKeown, Davide Martino, Janis M. Miyasaki, Michael G. Schlossmacher, A. Jon Stoessl, Antonio P. Strafella, Edward A. Fon, Oury Monchi

## Abstract

**Background:** Enhancing the interactions between study participants, clinicians, and investigators is imperative for advancing Parkinson’s disease (PD) research. The Canadian Open Parkinson Network (C-OPN) stands as a nationwide endeavor, connecting the PD community with ten accredited universities and movement disorders research centers spanning –at the time of this analysis– British Columbia, Alberta, Ontario, and Quebec.

**Objective:** Our aim is to showcase C-OPN as a paradigm for bolstering national collaboration to accelerate PD research and to provide an initial overview of already collected data sets.

**Methods:** The C-OPN database comprises de-identified data concerning demographics, symptoms and signs, treatment approaches, and standardized assessments. Additionally, it collects venous blood-derived biomaterials, such as for analyses of DNA, peripheral blood mononuclear cells (PBMC), and serum. Accessible to researchers, C-OPN resources are available through web-based data management systems for multi-center studies, including REDCap.

**Results:** As of November 2023, the C-OPN had enrolled 1,505 PD participants. The male-to-female ratio was 1.77:1, with 83% (n = 1098) residing in urban areas and 82% (n = 1084) having pursued post-secondary education. The average age at diagnosis was 60.2 ± 10.3 years. Herein, our analysis of the C-OPN PD cohort encompasses environmental factors, motor and non-motor symptoms, disease management, and regional differences among provinces. As of April 2024, 32 researchers have utilized C-OPN resources.

**Conclusions:** C-OPN represents a national platform promoting multidisciplinary and multisite research that focuses on PD to promote innovation, exploration of care models, and collaboration among Canadian scientists.

**PLAIN LANGUAGE SUMMARY:** Teamwork and communication between people living with Parkinson’s disease (PD), doctors, and research scientists is important for improving the lives of those living with this condition. The Canadian Open Parkinson Network (C-OPN) is a Canada-wide initiative, connecting the PD community with ten accredited universities and movement disorders research centers located in –at the time of this analysis– British Columbia, Alberta, Ontario, and Quebec. The aim of this paper is to showcase C-OPN as a useful resource for physician and research scientists studying PD in Canada and around the world, and to provide snapshot of already collected data. The C-OPN database comprises de-identified (meaning removal of any identifying information, such as name or date of birth) data concerning lifestyle, disease symptoms, treatments, and results from standardized tests. It also collects blood samples for further analysis.

As of November 2023, C-OPN had enrolled 1,505 PD participants across Canada. Most of the participants were male (64%), living in urban areas (83%), and completed post-secondary education (82%). The average age at diagnosis was 60.2 ± 10.3 years. In this paper, we look at environmental factors, motor and non-motor symptoms, different disease management strategies, and regional differences between provinces. In conclusion, C-OPN represents a national platform that encourages multidisciplinary and multisite research focusing on PD to promote innovation and collaboration among Canadian scientists.

## 1. INTRODUCTION

Parkinson’s disease (PD) is one of the fastest growing neurodegenerative conditions in terms of both prevalence and mortality [1,2]. Although symptomatic pharmacotherapy is available, no treatment stops the neuronal loss and clinical decline. Neuropathological hallmarks of PD comprise progressive nigrostriatal dopamine depletion and formation of a-synuclein-containing proteinaceous inclusions (Lewy bodies and neurites) [3]. Cardinal manifestations of PD include bradykinesia, rigidity, rest tremor, and postural instability. Non-motor features, such as hyposmia, rapid eye movement sleep behaviour disorder (RBD), constipation and other autonomic dysfunction, cognitive deficits, pain, depression, and anxiety, complete the clinical picture. The disease is much more heterogenous than previously appreciated in terms of clinical manifestations, etiopathogenesis, progression, and treatment response. Subgroups have been proposed based on clinical, genetic, and pathological features, but they remain poorly defined [4–7]. To gain a comprehensive understanding of these subgroups, it is essential to have large sample sizes and aggregate information.

The Canadian Open Parkinson Network (C-OPN; https://copn-rpco.ca/) is uniquely positioned to foster collaboration and facilitate the creation of extensive datasets and biosample collections on a national level, thereby encouraging the development of multisite and interdisciplinary partnerships. Launched in June 2020, the C-OPN bridges clinicians, researchers, and people with PD to facilitate innovative research in the Canadian and international landscapes. Given the complexity of PD, larger cohorts assembled in different geographical locations provide a high degree of biological and clinical diversity that benefits research on pathophysiological mechanisms, novel treatment strategies, and development of tools for prognostic, diagnostic, and disease management. The C-OPN holds the largest PD cohort in Canada with active recruitment sites in 11 Canadian cities (i.e., Vancouver, Calgary, Edmonton, London, Toronto, Ottawa, two in Montreal, two in Quebec City, and, recently, Halifax). The C-OPN also partners with the provincial Quebec Parkinson Network based at Montreal and Quebec City sites (QPN; [8]) as well as with the Calgary Parkinson Research Initiative (CaPRI). By doing so, the C-OPN is leveraged on the existence and expertise of established recruitment sites to expand its research promotion strategies to a national level. The C-OPN operates under Open Science and Open Data principles, meaning that data can also be leveraged, shared, and combined with other large-scale datasets [9].

Demographic, clinical, epidemiological, and cognitive assessment data are collected longitudinally. The network is uniquely positioned for facilitating research in that it also collects biological correlates from people living in Canada with PD, other atypical parkinsonism (AP), as well as non-neurological controls. The biological material collected consists of DNA, peripheral blood mononuclear cells (PBMCs), and serum. Further, the C-OPN registry links national and international researchers with participants for easier study recruitment. Interestingly, the C-OPN participants exhibit a 30% study response rate to such advertised studies. To date, C-OPN has recruited over 1,500 participants, with the PD subgroup being the largest.

Herein, we provide the methodology of C-OPN and present a description and a data analysis of the PD cohort specifically.

## 2. METHODS

### 2.1. Population and data collection

This study was approved by the Research Ethics Board of the University of Calgary (Calgary, AB; Ethics ID: REB19-1688_REN2). Only PD participants were included in the current analysis. Participants eligible for inclusion in the PD group were over 18 years of age and diagnosed by movement disorder specialists in Canada according to the Movement Disorder Society (MDS) criteria or previously published criteria such as the UK Brain Bank criteria [10,11]. To maximize recruitment and promote diversity, there are no exclusion criteria. Participants were recruited at various movement disorder clinics in four provinces across Canada: (i) the Pacific Parkinson’s Research Centre at the University of British Columbia (Vancouver, BC; Ethics ID: H19-01693); (ii) the Parkinson and Movement Disorders Program of the University of Alberta (Edmonton, AB; Ethics ID: Pro00091716_REN3); (iii) the Movement Disorders Program of the University of Calgary via the Calgary Parkinson Research Initiative (CaPRI) (Calgary, AB; Ethics ID: REB16-0545_REN6); (iv) the Parkinson Research Consortium of the University of Ottawa (Ottawa, ON; Ethics ID: 20190728-01H); (v) the Movement Disorders Centre at the University Health Network, University of Toronto (Toronto, ON; Ethics ID: 22-5071.0); (vi) the Movement Disorders Program of the University of Western Ontario (London, ON; Ethics ID: 2022-121756-74090); and four Movement Disorder Centres of the QPN, including (vii) the Montreal Neurological Institute (Montreal, QC; Ethics ID: IRB00010120); (viii) the Centre hospitalier de l’Université de Montréal (Montreal, QC; Ethics ID: F9H-92382); (ix) the CHU de Québec–Université Laval (Quebec, QC; Ethics ID: F9H-Distant-3720-92384); and (x) Clinique Neuro Lévis (Quebec, QC; Ethics ID: F9H-Distant-3720-92384). All participants provided written and informed consent, either in-person or electronically.

Data were collected using a panel of C-OPN questionnaires through the REDCap software, which includes the following instruments: (i) Enrollment; (ii) Demographic Questionnaire; (iii) Clinical Questionnaire; (iv) Medications Questionnaire; (v) Epidemiological Questionnaire; (vi) Montreal Cognitive Assessment (MoCA); and (vii) MDS-Sponsored Revision of the Unified Parkinson’s Disease Rating Scale (MDS-UPDRS). Key longitudinal data are collected from participants every 18 months for 3–5 years from enrollment, though only available for a small percentage due to recent recruitment. Questionnaires were completed virtually, in-person, or over the phone with assistance from a research coordinator or movement disorders specialist. Motor assessments (i.e., MDS-UPDRS) were conducted in-person by trained personnel, and blood draws from consenting participants were also performed during these visits. Cognitive assessment (i.e., MoCA-30 version 7.1) was completed either in-person or over the phone via T-MoCA (Ottawa site only), a modified version of the MoCA-30 version 7.1 [12], with trained personnel. All T-MoCA scores were converted to MoCA-30 scores according to methods described previously [12]. All data acquisition occurred directly into the REDCap version 13.7.31 database.

Blood samples from consenting participants in the four provinces were sent to the Montreal Neurological Institute (Montreal, QC) for processing (into DNA, PBMCs, and serum) and storage at the Clinical Biological Imaging and Genetic Repository (C-BIGR). Dopaminergic medication dosages were converted to levodopa (L-dopa) equivalent daily dose (LEDD) using an established formula (https://www.parkinsonmeasurement.org) [13].

### 2.2. Statistical analysis

The data supporting the findings of this study are available on request from the corresponding author. The data are not publicly available due to privacy or ethical restrictions.

Data are presented as percentages for categorical variables. Denominators for individual data fields were calculated based on the total number of responses recorded among the 1505 participants analyzed. As full neuropsychological evaluation was not available on all participants, we evaluated the theoretical prevalence of mild cognitive impairment (MCI) and dementia. For this purpose, we implemented the optimal screening cut-off point of 26 on the MOCA for MCI and the optimal diagnostic cut-point of 17 for dementia [14].

Quantitative variables were described with means and standard deviation and/or medians and interquartile ranges if non-normal distribution was confirmed by the Shapiro-Wilk’s test. Differences between patients recruited in eastern (Québec and Ontario) and western (Alberta and British Columbia) Canada were assessed using the Mann-Whitney U statistic, and global variations among provinces were evaluated with the Kruskal-Wallis test (correction for multiple comparisons implemented with the Dunn’s test and Sidak method). Fisher’s exact test was used for analyzing differences in qualitative variables. Data differing significantly between eastern and western Canada and within the four provinces were reported only if statistically similar values were observed among the pair of contiguous provinces from each region.

To address potential collinearity among significant variables, we conducted binary logistic regression to compare eastern and western Canada, and multinomial regression for inter-provincial analyses, excluding fields with ≥20% missing data. Regressors were selected using the stepwise backward method. In the Results section, we highlight variables differing significantly in at least 2 of the 6 inter-provincial comparisons.

Analyses were carried out in Stata 16 (StataCorp, College Station, USA). Choropleth maps were produced under the Albers projection using the GeoNames database from Microsoft Bing (Redmond, Microsoft Corporation, USA).

## 3. RESULTS

### 3.1. Enrollment in C-OPN

The workflow from participant recruitment to study participation is described in **Figure 1A**. A total of 1681 participants were included in the C-OPN database as of November 2023, which includes 1505 participants in the PD subgroup, 73 participants in the AP subgroup, and 103 participants in the control subgroup. For the PD subgroup, most participants (60%, n = 814) were recruited directly into C-OPN at various movement disorder clinics. Participants could also sign-up for C-OPN via the website (https://copn-rpco.ca/) (26%, n = 359), and the remaining participants were recruited by other means (14%, n = 169). The breakdown of site recruitment numbers across four Canadian provinces is depicted in **Figure 1B** and **Table 1**. Among those, 95% (n = 1430) are currently enrolled and 5% (n =75) have withdrawn for various reasons. Though the C-OPN was only recruiting in four provinces, some participants report living outside these provinces (**Figure 1C**). Table 1 provides a site breakdown of data and biosamples collected for the PD subgroup.

**Figure 1.**
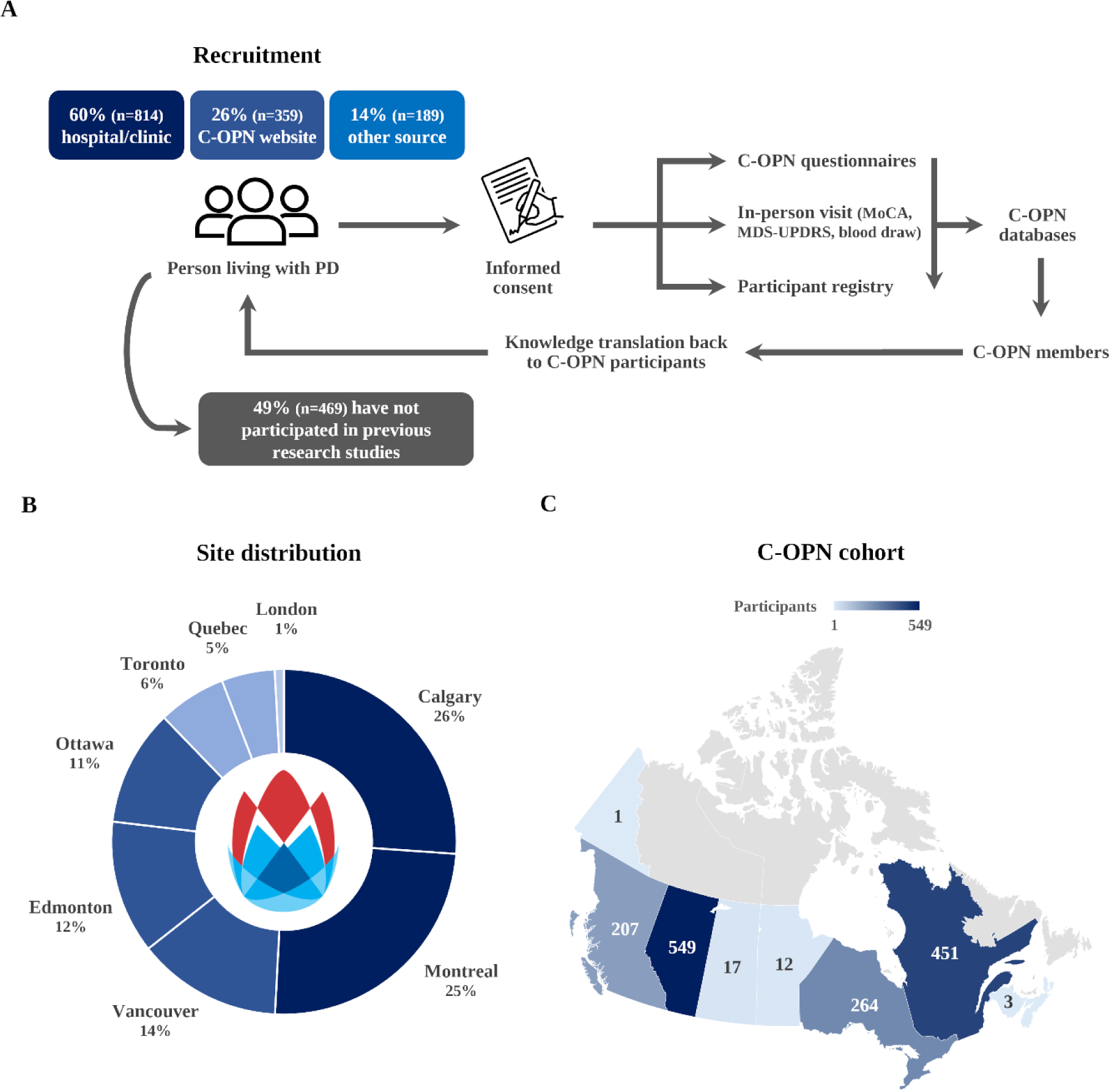
Breakdown of C-OPN workflow for collection of participant data and biosamples. Blue boxes, N = 1362; grey box, N = 957 **(A)**. Participant distribution by enrollment site **(B)** and location (postal code) **(C)**, N = 1505.

**Table 1.**
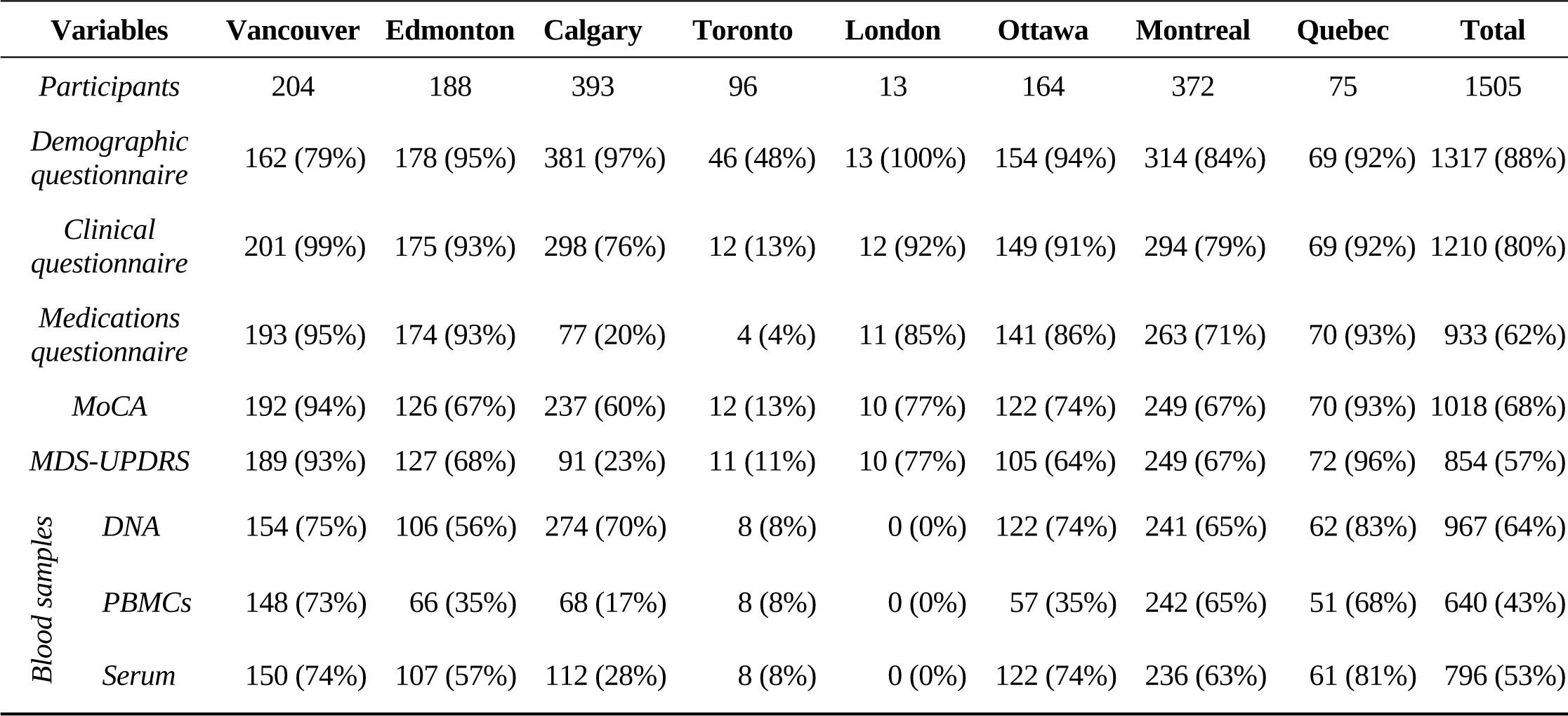
Data and biosample completion rate of C-OPN participants with PD. Data collection via REDCap software, variable N (640-1317).

### 3.2. Demographic characteristics of Participants with PD

At enrollment, the average age was 66.6 ± 9.4 years, and the median was 68 IQR 61-74 (n = 1281). The male: female ratio was 1.77:1 (36%, n = 502 females). Ninety-five percent (n = 1244) of participants reported Caucasian or French-Canadian ethnicity based on parental ethnicity. Of the participants, 82% (n = 1084) reported pursuing post-secondary education. Regarding living environments, 83% (n = 1098) resided in urban areas (population > 100,000 people), while 17% (n = 217) lived in rural areas (population < 100,000 people). Several comorbidities were reported among participants (**Figure. 2**).

**Figure 2.**
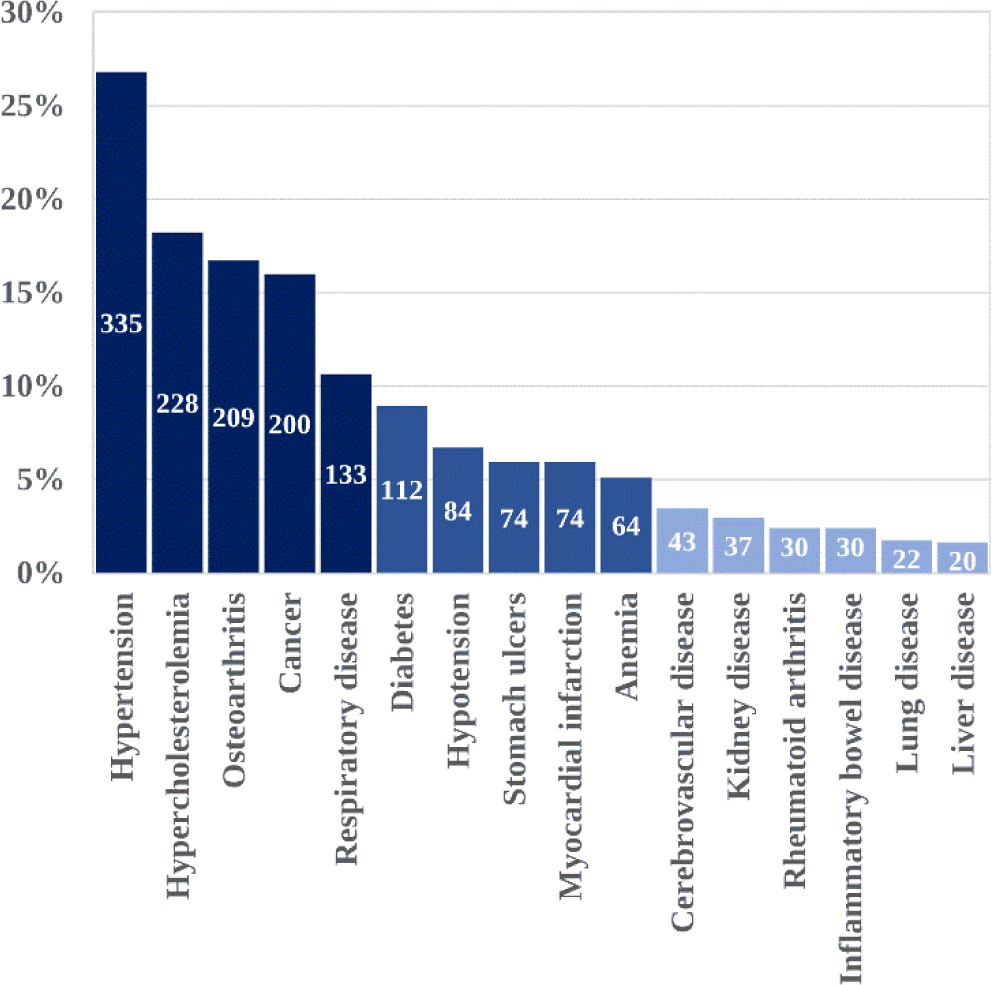
Comorbidities of C-OPN participants with PD. N = 1250.

### 3.3. Clinical characteristics of C-OPN participants with PD

The average age at diagnosis was 60.2 ± 10.3 years and the median was 61 IQR 53-68 (n = 1245), with the average duration since diagnosis being 8.4 ± 5.6 years and the median 7 years IQR 4-11 (n = 1245). Among the participants, 249 (31%) reported a family history of PD (**Figure 3**). The top reported first symptom (prior to diagnosis, at disease onset) (69%; n = 838) and current symptom (80%; n = 897) was tremor (**Figure 4A**, circles and bars respectively). Additionally, at the time of enrollment, 42% (n = 459) reported dyskinesia, 29% (n = 317) reported freezing, and 25% (n = 280) reported falling at least once in the three months prior (Figure 4A). As expected, symptom asymmetry was noted by most participants, with 42% (n = 414) reporting left-sided predominance and 47% (n = 458) reporting right-sided predominance. Only 9% (n = 86) reported symptoms affecting both sides equally and 2% (n = 21) reported undetermined symptom asymmetry. In general, 9% (n = 132) of participants were left-handed, 72% (n = 1010) were right-handed, and 2% (n = 15) were ambidextrous.

**Figure 3.**
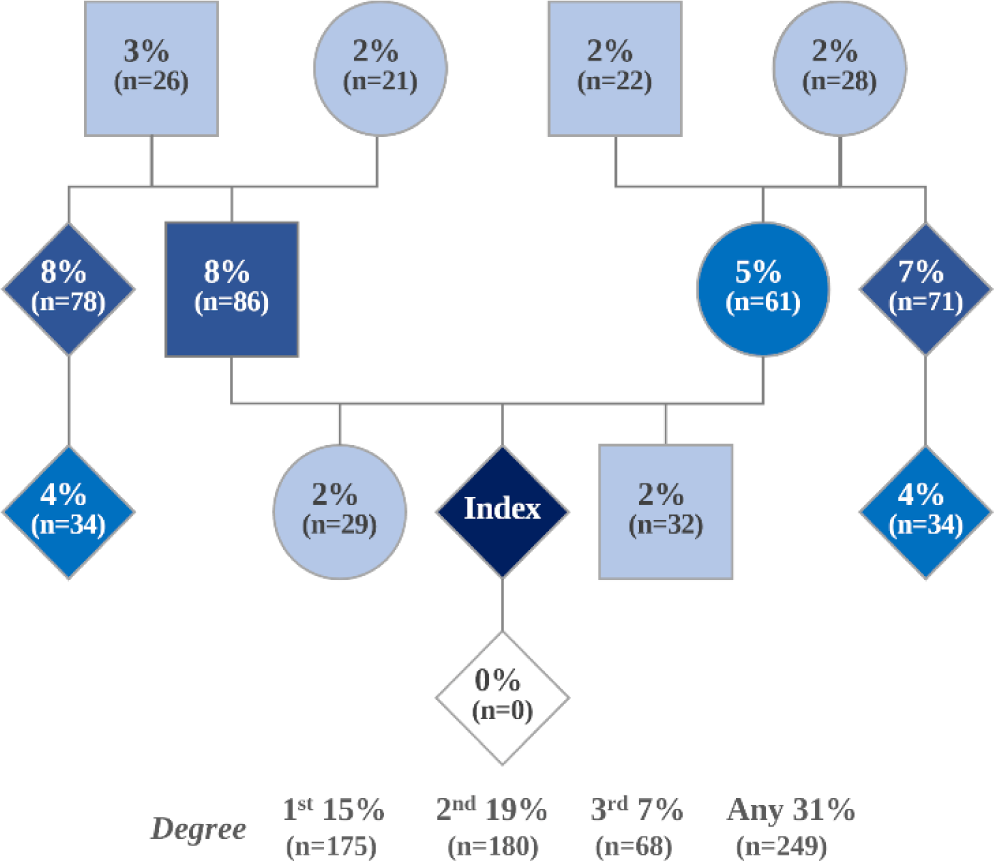
Family history of PD among C-OPN participants. Variable N (962-1247).

**Figure 4.**
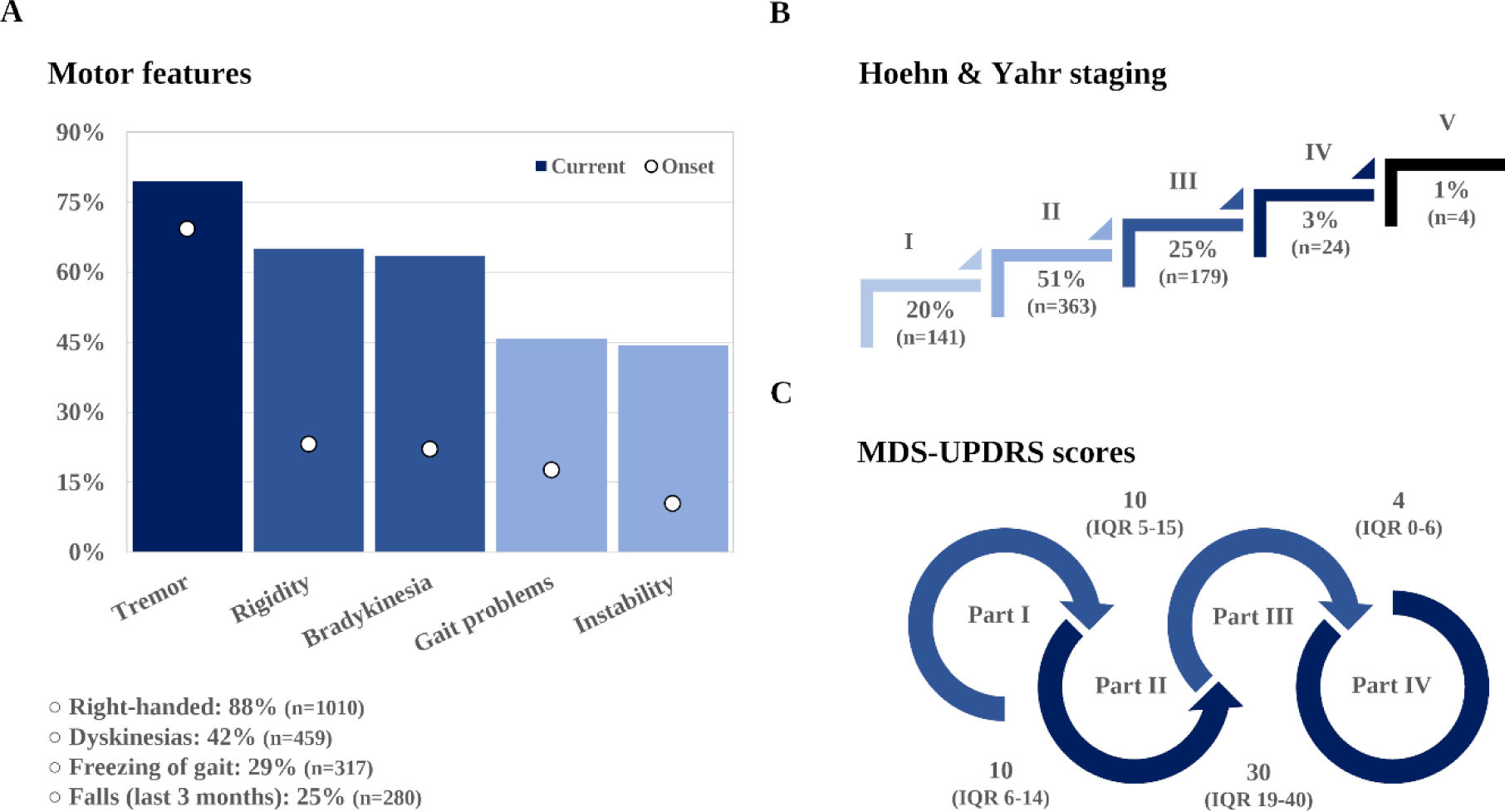
Cardinal motor features of PD. Top reported symptoms at onset and enrollment, N = 1210 and 1128, respectively; handedness, N = 1142; dyskinesia, N = 1081; freezing, N = 1087; falls, N = 1104 **(A)**; Hoehn & Yahr stage, N = 711 **(B)**; MDS-UPDRS score, N = 826 (part 1), N = 827 (part 2), N = 854 (part 3), and N = 818 (part 4) **(C)**.

The median Hoehn and Yahr (H&Y) score for the entire cohort was 2 IQR 2-3. A breakdown of scores by stages is depicted in **Figure 4B**. Despite delays in in-person visits caused by the COVID-19 pandemic, 797 complete MDS-UPDRS exams (89% performed in the on state) have been conducted thus far. The median score for each part is shown in **Figure 4C**.

In addition to motor signs and symptoms, the prevalence of common non-motor signs and symptoms in our cohort was assessed (**Figure 5A**). Average score on completed MoCA or T-MoCA Tests (n = 1018) was 25.8 ± 3.2, 26.5 IQR 24-28 (**Figure 5B**). Based on validated MoCA score cut-offs, 50% (n = 509) of participants screened positively for mild cognitive impairment (MCI; cut-off: ≤ 26 [14]), while 2% (n = 22) fell below the diagnostic cut-off for dementia (cut-off: ≤ 17 [14]). Further, 31% (n = 380) reported a close contact having noticed an increase in forgetfulness in the participant and 55% (n = 671) reported a decrease in their short-term memory. Regarding psychiatric features, 27% (n = 297) self-report anxiety, 27% (n = 299) depression, and 6% (n = 63) apathy. Additional hallmark non-motor symptoms, such as hyposmia, constipation, unexplained chronic pain, as well as sleep disturbances, are also reported in Figure 5A.

**Figure 5.**
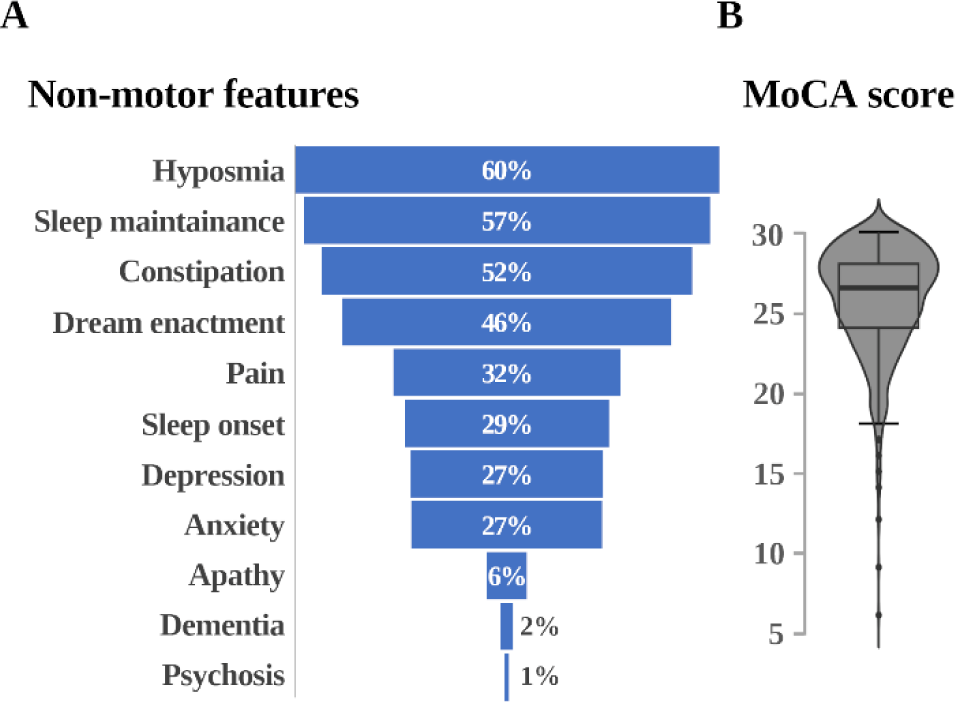
Common non-motor features of PD. Hyposmia, N = 1156; sleep disturbances, N = 1210 (difficulties falling asleep), N = 1178 (difficulties staying asleep), and N = 1187 (dream enacting behavior); constipation, N = 1247; pain, N = 1129; depression, N = 1109; anxiety, N = 1109; apathy, N = 1109; informed diagnosis of dementia, N = 1077; psychosis, N = 1109 **(A)**; MoCA, N = 1018 **(B)**.

### 3.4. Exposure to common environmental factors potentially contributing to PD

Common environmental factors that were correlated with PD in the literature were observed in the PD cohort before and/or after the diagnosis (**Figure 6**). Prior significant head trauma (i.e., concussion) was reported by 48% (n = 611) of participants, and 33% (n = 427) had previously engaged in high contact sports. Additionally, 51% (n = 668) of participants reported evident pesticide exposure, and 12% (n = 159) welding exposure. The type of contact sport, pesticide, and welding exposures were not collected. Seventy-two percent of participants (n = 960) reported current coffee consumption, while 63% (n = 822) reported drinking alcohol. Cigarette smoking was reported by 2% (n = 31) of participants, and 8% (n = 108) reported current cannabis consumption (legalized and regulated by the Canadian government in October 2018). Details on consumption can be found in Figure 6.

**Figure 6.**
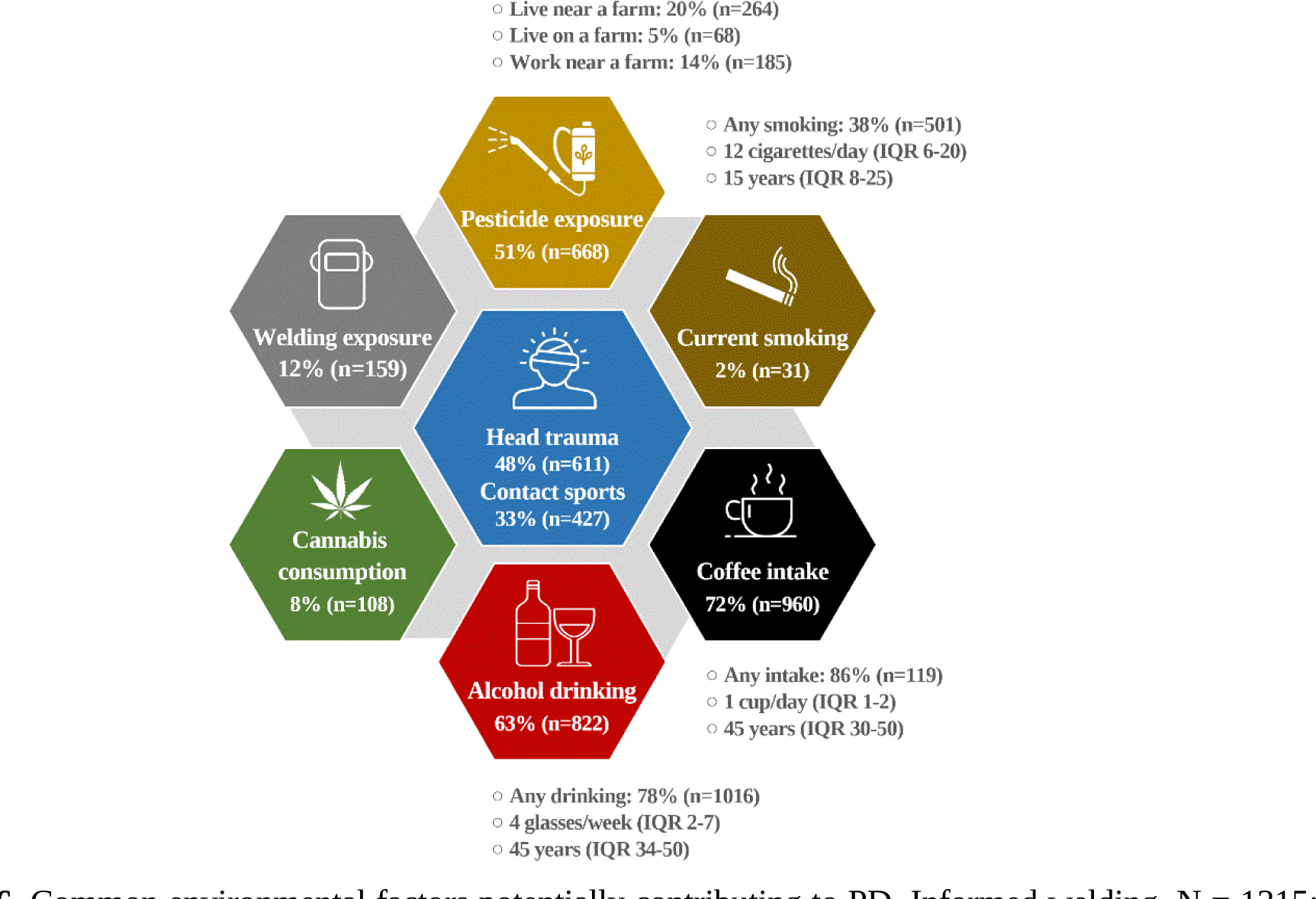
Common environmental factors potentially contributing to PD. Informed welding, N = 1315; informed pesticide exposure, N =1313; smoking, N = 1313; coffee, N = 1316; alcohol, N= 1309; cannabis, N = 1309; head trauma, N = 1265 (serious head injury) and N = 1294 (contact sports).

### 3.5. Non-clinical and clinical management of PD in Canada

Non-pharmacologic management of PD varies widely. Only 39% (n = 423) have regular caregivers, primarily spouses (92%, n = 389). Additionally, 23% (n = 245) participate in support groups. Physical activity is a major approach, with 77% (n = 880) exercising regularly, averaging 5.5 ± 3.8 times per week for at least 30 minutes. Top activities include walking, biking, and weight training.

Regarding dopaminergic medications, the average LEDD was 799.13 ± 549.90 mg, with a median of 688 mg IQR 450-1050. Among those taking dopaminergic medications, 91% (n = 840) reported symptom improvement, while 9% (n = 82) reported lack of it. Dopamine agonist use included pramipexole (16%, n = 178), rotigotine (4%, n = 41), ropinirole (1%, n = 14), apomorphine (0.2%, n = 2). Further, 16% of participants took a dopamine agonist in combination with levodopa. Other medications for motor symptoms included amantadine (17%, n = 155), botulinum toxin injections (2%, n = 18), anticholinergics (2%, n = 14), and other PD medications (0.7%, n = 6).

Considering that non-motor symptoms are common amongst PD patients, several adjuvant medications can also be prescribed. These includes: (i) antidepressants (20%, n = 271); (ii) analgesics (9%, n = 119); (iii) benzodiazepines (5%, n = 73); (iv) antipsychotics (2%, n = 30); (v) cholinesterase inhibitors such as donepezil or rivastigmine (2%, n = 21); (vi) stimulants (0.4%, n = 5); or (vii) other types of non-PD medications not previously listed (63%, n = 846). While some participants listed melatonin as part of their medication regime, there was no question specifically targeting this supplement.

### 3.6. Differences by province and region

The C-OPN provides a unique opportunity to compare data collected between provinces spanning eastern and western Canada (**Figure 7**). Significant global variations were observed for several factors when comparing British Columbia, Alberta, Ontario, and Quebec, including age at PD onset (p = 0.015), age at enrollment (p = 0.021), family history of PD in paternal uncles/aunts (p = 0.043), proximity to farms (p = 0.013), initial symptoms like instability (p = 0.049), and unexplained pain (p = 0.021). Variations were also noted in medication prescriptions, including levodopa formulations (p < 0.050), pramipexole (p < 0.001), anticholinergics (p = 0.037), and botulinum toxin (p < 0.001). Access to therapies such as physical (p = 0.018), speech (p = 0.015), and swallowing (p = 0.005), as well as surgical interventions like DBS (p < 0.001), differed between provinces.

**Figure 7.**
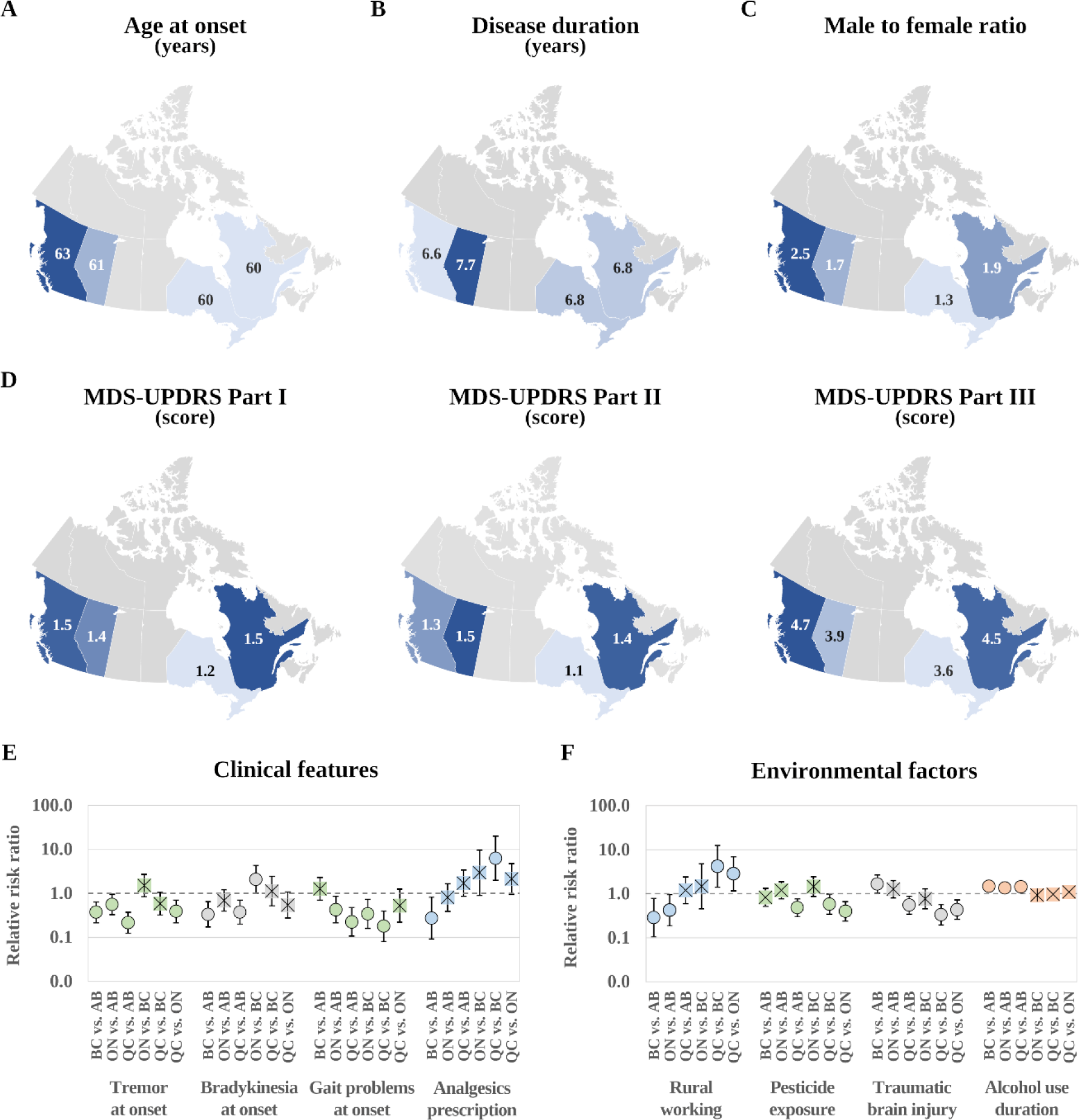
Key features of C-OPN participants with PD in the four most populous Canadian provinces. Description by province of age at onset (**A**), disease duration (**B**), male to female ratio (**C**), and MDS-UPDRS parts 1-3 (**D**). Multinomial comparisons by clinical features (**E**), and key environmental factors (**F**); **w**ithin the forest plots, significant comparisons depicted with a circle (o) are significant (*p <* 0.050), while those with a crossed square (☒) are not.

Further analysis through multinomial regression (Figure 7E-F) revealed additional insights. Participants from Alberta reported reduced alcohol intake duration. Alberta and Quebec had more participants living and working in rural settings, with Quebec reporting the lowest exposure to pesticides and head trauma. Quebec and Ontario participants tended to exhibit fewer tremors, bradykinesia, and gait problems at disease onset. Conversely, PD participants from British Columbia showed lower analgesic use compared to those from Alberta and Quebec.

Additionally, Western subjects experienced older onset (OR 1.23 CI95% 1.08-1.39, p = 0.002) and had higher odds of receiving subthalamic DBS (OR 6.28 CI95% 2.12-18.57, p = 0.001). History of PD in brothers was less common among Western patients (OR 0.37 CI95% 0.16-0.87). Complete models are available in the Supplementary Material (**Table S1**).

## 4. DISCUSSION

C-OPN collects a vast amount of clinical information, enabling it to provide robust, biologically informed diagnostic criteria for personalized treatment strategies, to contribute to the development of more accurate prognosis tools, and to the stratification of cohorts for trials of disease-modifying therapies. Further, in addition to people with PD, C-OPN also recruits people living with other parkinsonian syndromes (including progressive supranuclear palsy, multiple system atrophy, corticobasal syndrome, dementia with Lewy bodies, frontotemporal dementia, etc.) as well as non-neurological controls, which comprise important comparison populations to evaluate the specificity of an effect on PD. To date, 32 projects and counting have utilized C-OPN resources to facilitate their research efforts, including both national and international scientists [15,16].

The α-synuclein seed amplification assay findings from the Michael J. Fox Foundation (MJFF), published in April 2023 during Parkinson’s Awareness Month, would not have been feasible without a national, multi-site, and collaborative research initiative, such as Parkinson’s Progression Marker Initiative (PPMI). Like PPMI, C-OPN fits this role within the Canadian PD landscape. Similar initiatives in Canada – such as the Ontario Neurodegenerative Disease Research Initiative (ONDRI) [17] led by the Ontario Brain Institute (OBI), and the Comprehensive Assessment of Neurodegeneration and Dementia (COMPASS-ND) study [18] led by the Canadian Consortium on Neurodegeneration in Aging (CCNA), as well as PPMI in select Canadian cities –have likewise established cohorts to better define neurodegenerative diseases and their many subtypes. What makes C-OPN unique compared to the above-mentioned cohorts is (i) the collection of PBMCs, a highly useful tool in clinical research and drug development; (ii) the scale at which participants are being recruited across Canada, as C-OPN represents the largest parkinsonian cohort in the country; and (iii) the ability to compare and contrast demographic profiles and disease management strategies by region, particularly as C-OPN continues to expand across the country.

Stratification of distinct PD phenotypes in clinical trials, similar to analyses in the PPMI study, may allow for a better understanding of why some patients respond to treatment, while others show no effect. This would be a game-changer for people living with PD or related disorders, and lead to significant improvements in quality of life and the potential for disease-modifying interventions. Preliminary analyses demonstrate that the C-OPN cohort is a representative PD cohort from tertiary and academic centers, comparable to previous reports describing different clinical and epidemiological characteristics and correlations within similar PD cohorts. In addition to the PPMI led by MJFF [19], this also includes the French clinical research network for PD (NS-Park) [20], the Harvard Biomarkers Study [21], the Luxembourg Parkinson’s study [22], COURAGE-PD [23], the UK Biobank [24], and the DeNoPa cohort in Germany [25], among others. For example, C-OPN’s PD cohort displays epidemiological characteristics similar to NS-Park’s previously published cohort, including age at disease onset (58.5 ± 11.4 years), disease duration (9.2 ± 6.9 years), and sex distribution (1.4:1), among others [20].

Selection biases in the cohort include overrepresentation of participants of Caucasian ethnicity and higher education levels compared to the general population. Characteristics related to disease onset, motor and non-motor symptoms, and management strategies align with other PD cohorts, with notable findings and differences outlined below. A shortcoming of our current enrolment structure is the paucity of control individuals, which we seek to correct in the coming years. Sampling of persons with other neurological diseases as well as those without any disorder of the nervous system may be important for reducing bias in future analyses.

### 4.1. Clinical characteristics of PD

The most prominent first symptom reported (prior to receiving a diagnosis of PD) was tremor. Interestingly, postural instability and gait problems were also reported as initial symptoms by 10% and 18% of participants, respectively. Symptoms relating to balance and gait are typical early manifestations of other forms of parkinsonism, such as progressive supranuclear palsy, and could suggest possible misdiagnosis. Particularly during early stages of the disease, misdiagnosis of Parkinsonism can be a significant concern due to overlap in symptom profiles between Parkinsonian syndromes [26].

Moreover, there are roughly 770 community neurologists in Canada and fewer than 80 neurologists specialized in treating movement disorders, with most of these specialists located in major urban centres within the provinces [27]. This poses a significant geographical barrier for people living with PD in rural, remote, and northern communities across the country. One way C-OPN is working to address this gap is by including more people living in these communities in research studies via an online/at-home model of the already existing C-OPN infrastructure, set to be launched in the coming year.

The most reported clinical manifestations encompass the cardinal motor signs and symptoms of PD. These includes tremor, rigidity, bradykinesia, postural instability, gait problems, dyskinesia, freezing, and falling. With respect to postural instability, this self-reported statistic is interesting, considering it was reported as a current symptom by nearly half of all participants (44%), while the median H&Y score of the cohort is 2. Mild postural imbalance is not typically seen until H&Y stage 3. Regarding symptom asymmetry, early symptoms were reported to affect either the left or right side equally, despite most participants being right-hand dominant (like the general population). This suggests that symptom asymmetry is not influenced by handedness. The top-reported comorbidities were hypertension (27%), hypercholesterolemia (18%), and osteoarthritis (17%). These rates are similar in range to those in the general population of Canada [28–30]: hypertension (25%), hypercholesterolemia (28%), and osteoarthritis (14%).

When comparing between Eastern (Ontario and Quebec) and Western (Alberta and British Columbia) provinces, motor symptoms varied at onset. Eastern provinces had a lower proportion of tremor (particularly Quebec), bradykinesia, and problems with gait at disease onset. Though further analysis into these differences is beyond the scope of the current manuscript, future studies may explore these differences and how they might influence province-specific management practices of PD. This will also be an important step in better understanding access to care across Canada as it relates to PD, especially considering the responsibility to administer and deliver most of Canada’s healthcare services lies within the provinces and territories.

Many of the common non-motor symptoms of PD were reported in the C-OPN PD cohort, including memory problems and cognitive decline, sleep disturbances, hyposmia, constipation, pain, anxious, and depressive mood. Although apathy, another frequently observed non-motor symptom of PD, was not reported as widely, it is important to note that a dedicated apathy evaluation, like the Lille Apathy Rating Scale [31], was not conducted, which might have yielded a more representative result. Additionally, participants experiencing significant apathy may be less inclined to engage in research studies or may be less likely to self-report feelings of apathy overall. Further, in our enrolment protocol for baseline assessments, standardized quantification of olfaction has not yet been, but will be, included [18]. Although hyposmia is frequently reported subjectively, as we have registered in the C-OPN cohort, its prevalence is significantly higher when tested objectively in persons with PD. Its analysis may offer important clues to the start of PD as well as to distinct subtypes and co-pathologies of neurodegeneration [32,33].

### 4.2. Management of PD

A wide spectrum of management practices exists as part of the treatment strategy of PD. Exercise is among the most widely studied and shown to improve many symptoms of PD, including balance, gait, risk of falls, physical function, sleep impairments, cognitive function, and quality of life [34]. Effective exercise interventions include gait and balance training, progressive resistance training, treadmill exercise, strength training, aerobic exercise, music- and dance-based approached, and tai chi [35], many of which were verbally reported by C-OPN participants.

The diverse combination of motor and non-motor features of PD have led to the development of subtyping approaches, whereby different subtypes of PD may respond differently to various anti-PD medications. One of these approaches proposes the presence of three subtypes, which comprise: (i) mild motor predominant PD, characterized by a younger age at onset, mild motor and non-motor symptoms, slow progression, and good medication response; (ii) intermediate PD, characterized by intermediate age at onset and symptomatology, moderate-to-good response to medications; and (iii) diffuse malignant PD, most notably characterized by baseline motor symptoms accompanied by rapid eye movement sleep behaviour disorder, mild cognitive impairment, orthostatic hypertension, worse levodopa response, more prominent dopaminergic dysfunction on DaT SPECT, more atrophy in specific brain regions, low amyloid-β and τ:amyloid-β ratio in the cerebrospinal fluid, and rapid progression [35]. Considering that 82% of C-OPN participants reported a positive response to initiation of a dopaminergic agent, it is likely that most C-OPN participants with PD comprised the first two subtypes, even though an assessment of imaging and cerebrospinal fluid markers was not available from the larger cohort. In the absence of these biomarkers, it is difficult to comment on the prevalence of the more severe subtype but note that many of our participants were still in relatively early stages of disease and the scientific community has yet to agree on a subtyping approach.

### 4.3. Genomics

The C-OPN is a member of the Global Parkinson’s Genetics Program (GP2), a global collaboration project of the Aligning Science Across Parkinson’s (ASAP) initiative focused on improving our understanding of the genetic architecture of PD and making this knowledge globally relevant. DNA samples from all C-OPN participants are being sent to GP2 for genome-wide association studies (GWAS). To date, DNA samples from > 494 participants have been sent to GP2, among which 127 of these participants reported having a family history of PD –with the majority citing first- or second-degree relatives– and these select DNA samples will undergo whole genome sequencing as part of the GP2 monogenetic hub project. Results from these genetic analyses will be continuously added to an existing collection of genetic data for PD patients via C-BIGR, the largest of its kind available in Canada, all of which is made available to C-OPN members.

### 4.4. Navigating Open Science and Open Data in Canada

C-OPN has continued to adapt and find innovative solutions to practicing under Open Science principals in Canada, while consistently upholding the highest level of data security. Coordinating and obtaining ethics approval and data/material transfer agreements at each of the sites across Canada proved to be an arduous task. This is, in part, due to the lack of a national, unified, and standardized ethics review board in Canada. Nevertheless, since its launch, C-OPN has successfully obtained ethics board approval and implemented appropriate legal agreement at 11 sites in five provinces across Canada, with more sites projected to join in the future. In fact, in March 2024, the Halifax C-OPN site, our first Atlantic Canada site, recently received ethics board approval via Centricity Research, was launched, and is now recruiting participants.

### 4.5. Major limitations

As C-OPN expands nationwide, prioritizing outreach to remote, northern, and Indigenous communities becomes crucial, given their historical underrepresentation in PD research and other fields. Despite limitations in data collection methods, C-OPN continuously enhances online questionnaires, adjusting language and scope for more meaningful and representative data collection. However, some variables lack validation or quantification; for instance, data on psychiatric symptoms such as apathy, anxious, and depressive mood were gathered without validated questionnaires. Moreover, specific information on antipsychotic or antidepressant medications was not collected. As more networks emerge addressing various neurological conditions, including PD, ensuring compatibility between organizations may pose a challenge, yet remains an important objective.

### 4.6. Conclusions

Studying the phenotypic variability of PD and its contributors is crucial for personalized treatment options to be developed. C-OPN addresses this gap by providing access to diverse and large-scale data sets, aiding in understanding PD’s pathophysiology and variable trajectories. A critical next step for C-OPN is to overcome barriers to subject enrollment across diverse provinces with heterogenous populations, which is highly relevant for a complex disease like PD. Ultimately, C-OPN aims to support and expedite research, facilitating early access to disease-modifying interventions for all patients with parkinsonism.

## Data Availability

All data produced in the present study are available upon reasonable request to the authors.

## ACKNOWLEDGEMENTS

We thank the participants in C-OPN and their families.

## 7. AUTHOR CONTRIBUTIONS

MC, GPM, MB, CPN contributed to this manuscript equally, with MC writing the first draft. GDPM led the data analysis. All co-authors contributed to the revision of this manuscript.

---

**Table S1.**
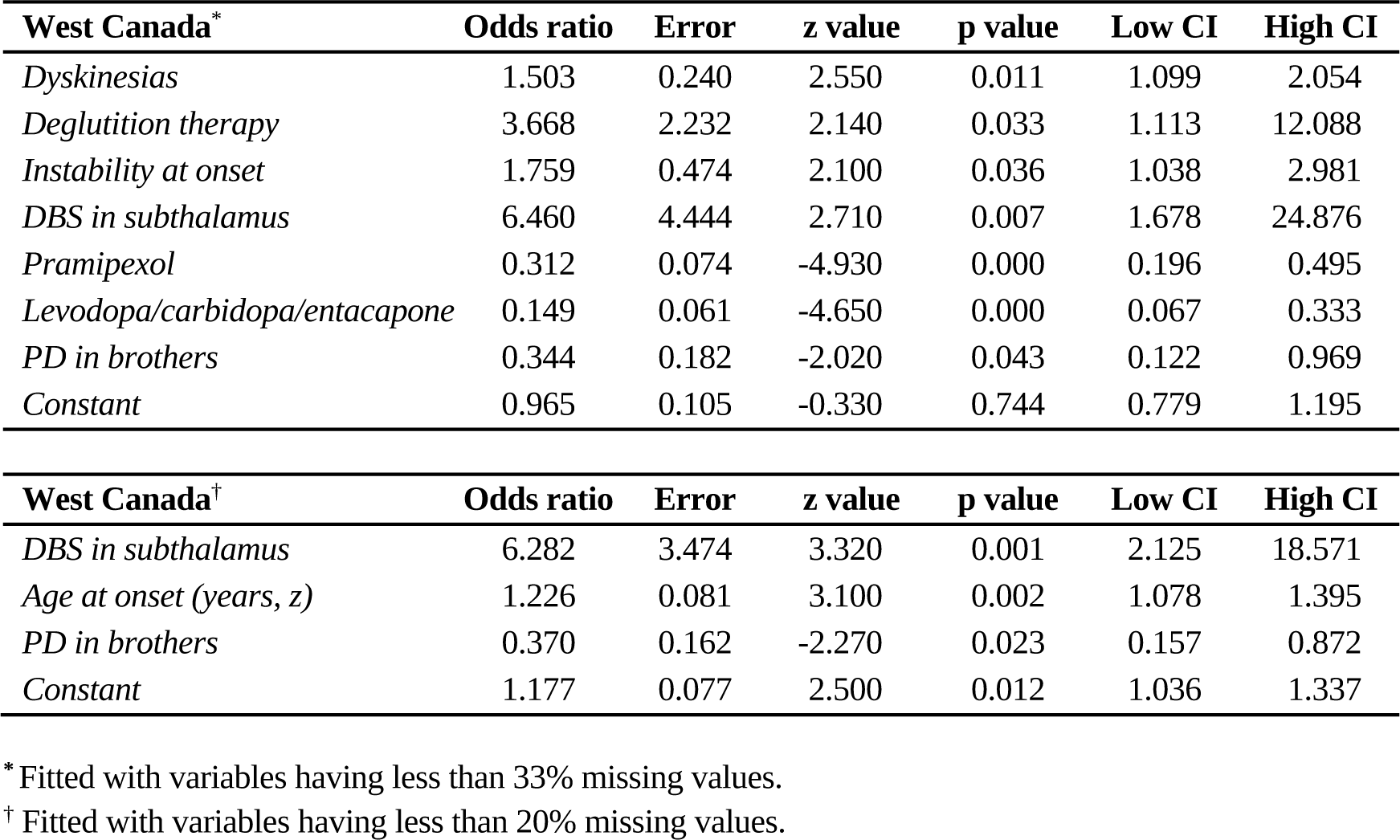
Differences in key features by region. Western Canada (British Columbia and Alberta) relative to eastern Canada (Ontario and Quebec).

| **West Canada\*** | **Odds ratio** | **Error** | **z value** | **p value** | **Low CI** | **High CI** |
| --- | --- | --- | --- | --- | --- | --- |
| *Dyskinesias* | 1.503 | 0.240 | 2.550 | 0.011 | 1.099 | 2.054 |
| *Deglutition therapy* | 3.668 | 2.232 | 2.140 | 0.033 | 1.113 | 12.088 |
| *Instability at onset* | 1.759 | 0.474 | 2.100 | 0.036 | 1.038 | 2.981 |
| *DBS in subthalamus* | 6.460 | 4.444 | 2.710 | 0.007 | 1.678 | 24.876 |
| *Pramipexol* | 0.312 | 0.074 | -4.930 | 0.000 | 0.196 | 0.495 |
| *Levodopa/carbidopa/entacapone* | 0.149 | 0.061 | -4.650 | 0.000 | 0.067 | 0.333 |
| *PD in brothers* | 0.344 | 0.182 | -2.020 | 0.043 | 0.122 | 0.969 |
| *Constant* | 0.965 | 0.105 | -0.330 | 0.744 | 0.779 | 1.195 |

| **West Canada†** | **Odds ratio** | **Error** | **z value** | **p value** | **Low CI** | **High CI** |
| --- | --- | --- | --- | --- | --- | --- |
| *DBS in subthalamus* | 6.282 | 3.474 | 3.320 | 0.001 | 2.125 | 18.571 |
| *Age at onset (years, z)* | 1.226 | 0.081 | 3.100 | 0.002 | 1.078 | 1.395 |
| *PD in brothers* | 0.370 | 0.162 | -2.270 | 0.023 | 0.157 | 0.872 |
| *Constant* | 1.177 | 0.077 | 2.500 | 0.012 | 1.036 | 1.337 |

\* Fitted with variables having less than 33% missing values.
† Fitted with variables having less than 20% missing values.

